# Cortical thickness and formal thought disorder in schizophrenia: An ultra high-field network-based morphometry study

**DOI:** 10.1101/2019.12.17.19014415

**Authors:** Lena Palaniyappan, Ali Al-Radaideh, Penny A. Gowland, Peter F. Liddle

**Affiliations:** Robarts Research Institute, University of Western Ontario, London, Ontario, Canada; Department of Psychiatry, University of Western Ontario, London, Ontario, Canada; Lawson Health Research Institute, London, Ontario, Canada; Department of Medical Imaging, Faculty of Allied Health Sciences, The Hashemite University, Zarqa, Jordan; Sir Peter Mansfield Imaging Centre (SPMIC), School of Physics and Astronomy, University of Nottingham, Nottingham, UK; Translational Neuroimaging for Mental Health, Division of Psychiatry and Applied Psychology, University of Nottingham, Nottingham, UK

**Author notes:** **Address** correspondence to: Dr. Lena Palaniyappan, Room-A2/636, Program for Prevention and Early Intervention in Psychoses, Victoria Hospital, 800, Commissioners Road, London, ON. Canada.

**Keywords:** Disorganisation, thought disorder, Salience Network, Cognitive Control, Language Network, coherence

## Abstract

**Background:** Persistent formal thought disorder (FTD) is a core feature of schizophrenia. Recent cognitive and neuroimaging studies indicate a distinct mechanistic pathway underlying the persistent positive FTD (pFTD or disorganized thinking), though its structural determinants are still elusive. Using network-based cortical thickness estimates from ultra-high field 7-Tesla Magnetic Resonance Imaging (7T MRI), we investigated the structural correlates of pFTD.

**Methods:** We obtained speech samples and 7T MRI anatomical scans from medicated clinically stable patients with schizophrenia (n=19) and healthy controls (n=20). Network-based morphometry was used to estimate the mean cortical thickness of 17 functional networks covering the entire cortical surface from each subject. We also quantified the vertexwise variability of thickness within each network to quantify the spatial coherence of the 17 networks, estimated patients vs. controls differences, and related the thickness of the affected networks to the severity of pFTD.

**Results:** Patients had reduced thickness of the frontoparietal and default mode networks, and reduced spatial coherence affecting the salience and the frontoparietal control network. A higher burden of positive FTD related to reduced frontoparietal thickness and reduced spatial coherence of the salience network. The presence of positive FTD, but not its severity, related to the reduced thickness of the language network comprising of the superior temporal cortex.

**Conclusions:** These results suggest that cortical thickness of both cognitive control and language networks underlie the positive FTD in schizophrenia. The structural integrity of cognitive control networks is a critical determinant of the expressed severity of persistent FTD in schizophrenia.

## Introduction

Formal thought disorder (FTD) is a core clinical feature of schizophrenia (Jerónimo et al., 2018). Presence of FTD predicts transition to psychosis in individuals at high risk of schizophrenia, predating the actual onset by several years (Bearden et al., 2011; Demjaha et al., 2017; DeVylder et al., 2014; Gooding et al., 2013; Wilcox et al., 2014). FTD is also indicative of higher illness severity (Roche et al., 2015) and poor socio-occupational outcomes (Roche et al., 2016) in first episode psychosis. While FTD seen in acute psychosis often resolves with time, persistence of FTD in those with established illness affects therapeutic alliance (Cavelti et al., 2016), social functioning (Palaniyappan et al., 2019b; Sousa et al., 2019) and occupational recovery (Harrow and Marengo, 1986), thus reducing the overall quality of life (Tan et al., 2014; Ulas et al., 2008). Despite this, our current understanding of the mechanistic basis of FTD is limited and treatment approaches are elusive (Kircher et al., 2018).

Two recent state-of-art syntheses in FTD highlight the substantial inconsistencies in the neuroimaging observations reported to date (Cavelti et al., 2018a; Sumner et al., 2018a). While functional MRI studies often employ linguistic tasks that preferentially implicate language network in FTD (Cavelti et al., 2018a; Sumner et al., 2018b; Wensing et al., 2017), structural MRI studies report more distributed abnormalities in both the language network and non-language specific networks (Cavelti et al., 2018a; Sumner et al., 2018a). In particular, the two distinct domains of FTD – positive FTD (characterised by looseness, peculiar word/sentence or illogicality) and negative FTD (characterised by reduced fluency, speed and content) – are thought to have distinct neurocognitive correlates (Bora et al., 2019; Tan and Rossell, 2017) but are not often parsed in neuroimaging studies. A small number of studies that parse these 2 dimensions identify structural deficits that extend beyond the language network (Palaniyappan et al., 2015; Sans-Sansa et al., 2013) in negative FTD. Nevertheless, the structural basis of positive FTD continues to be elusive in most studies (Cavelti et al., 2018b; Palaniyappan et al., 2015; Winkelbeiner et al., 2018; but see Sans-Sansa et al., 2013).

The mechanistic basis of positive FTD is particularly challenging to elucidate as the clinical features are often subtle (Cohen et al., 2017; Sommer et al., 2010), fluctuate with acute psychotic symptoms (Li et al., 2019) and easily missed during cross-sectional clinical interviews (de Bruin et al., 2007; Palaniyappan, 2009). Furthermore, most morphometric studies in FTD quantify the composite measure of grey matter volume (except (Winkelbeiner et al., 2018)). Cortical thickness is much more sensitive morphological metric that provides insights into dynamic structural remodeling related to longitudinal changes in symptom trajectories (Guo et al., 2016; Palaniyappan et al., 2019a)). While volume changes may result from non-specific morphometric effects (Palaniyappan and Liddle, 2012; Panizzon et al., 2009), thickness changes are increasingly linked to intracortical myelination and dendritic spine branching (Shin et al., 2018; Storsve et al., 2014), thus enabling us to hone in on cellular processes underlying the suspected structural changes in FTD (Kircher et al., 2018). There are no whole brain studies relating cortical thickness to FTD to date.

The thickness of cerebral cortex varies across individuals as well as across different cortical regions within an individual. Across subjects, the cortical thickness of one region of a functional network co-varies tightly with other regions that constitute the functional network possibly due to shared trophic influences and/or coordinated maturational programming (Alexander-Bloch et al., 2013b, 2013a). Spatially distributed regions within a defined functional network show consistent task-related recruitment, which can also have a plastic influence on the observed structural covariance (Kelly and Castellanos, 2014). At an individual level, pathological processes disrupting this maturational or functional coherence can result in higher than expected degree of variability within large-scale networks (Kong et al., 2019, 2014), resulting in *spatial incoherence*. Interestingly, patients with schizophrenia show both relative thinning and thickening of cortex when compared to controls, in functionally linked brain regions (e.g. thinning of posterior cingulate, but thickening of precuneus, within the same default-mode network as shown by van Erp et al., 2018), which can result in increased spatial incoherence within functional networks. We quantified the network-level thickness and its spatial incoherence in 17 cortical networks covering the entire cortical surface, identified on the basis of well-replicated patterns of intrinsic connectivity (Yeo et al., 2011).

The aim of the present study was to investigate if the severity of positive FTD varies with illness-related changes in cortical thickness in schizophrenia. To this end we utilised the same sample previously reported (in Palaniyappan et al., 2015) to first delineate the cortical networks showing significant thinning and spatial incoherence in patients with schizophrenia compared to healthy controls. We then studied if positive FTD was associated with the thickness and coherence of the affected cortical networks. Given the prior findings implicating language networks in FTD (Kircher et al., 2018), we investigated if structural changes in language networks showing intrinsic functional connectivity independently explain the variance in the severity of positive FTD, in addition to the illness-related morphometric changes.

## Methods

### Participants

20 patients receiving community based care for a Diagnostic and Statistical Manual of Mental Disorders (DSM)-IV (American Psychiatric Association, 1994) diagnosis of schizophrenia were specifically recruited in a stable phase of illness for this study. 21 healthy controls group matched for age, gender and parental socioeconomic status were also recruited. 1 patient and 1 control were excluded due to motion artifacts affecting cortical surface reconstruction. Subjects with neurological disorders, current substance dependence, or intelligence quotient (IQ) < 70 (Ammons and Ammons, 1962) were excluded. Healthy subjects were also excluded if there was a personal or family history of psychosis. All subjects were recruited from the county of Nottinghamshire, UK with informed consent to participate obtained in accordance with the approval from the National Research Ethics Committee, Nottinghamshire.

### Clinical Assessment

The diagnosis was made through a consensus procedure as per Leckman et al., (1982), using multiple sources of information including clinical notes, information provided by treating clinicians as well a structured diagnostic interview (Liddle et al., 2002). The clinical stability was assessed as <10 points change in Global Assessment of Function [GAF, DSM-IV (American Psychiatric Association, 1994)] score in the 6 weeks prior to study participation.

Patents and healthy controls were interviewed on the same day as the scan using the validated procedure for administering Thought Language Index (Liddle et al., 2002). Subjects were asked to describe 3 pictures from Thematic Apperception Test (Murray, 1943), for one minute each, with follow-up questions raised by the interviewer if needed. The free speech samples were recorded and later transcribed by a team member with no direct contact with the subject and blind to the diagnostic status, symptom burden and neuroimaging findings. Positive FTD was defined as the summed score of looseness, peculiar word, peculiar sentence and peculiar logic were classified as positive FTD while the scores of perseveration, poverty of speech and weakening of goal were summed as negative FTD, as in our previous study (Palaniyappan et al., 2015). All patients in our sample had English as their first language.

Handedness was assessed using the 12-items Annett scale (Annett, 1970). The median defined daily dose (DDD) of antipsychotics (WHO Collaborating Centre for Drug Statistics and Methodology, 2003) was calculated for all patients.

### MRI acquisition and processing

The procedures for MRI acquisition are described in our previous report (Palaniyappan et al., 2015). In summary, T1 weighted images were acquired from a 7T Philips Achieva (32-channel coil) using a 3D Magnetization Prepared-Turbo Field Echo (IR-TFE) with 0.6 mm isotropic resolution, 192 × 180 × 140 mm matrix, TR = 15 ms, TE = 5.6 ms, shot interval = 3 s, and flip angle 8°. Bias field inhomogeneity was reduced using an optimized inversion pulse (adiabatic pulse). We have previously shown that in this sample, the total grey matter tissue volume estimated from the 3T and 7T scans from the same subjects were not different (paired t test t=0.18, *p*=0.9) and were very highly correlated (r = 0.93, *p*<.001), indicating that there were no systematic differences in the tissue segmentation of 7T scans due to bias inhomogeneity (Iwabuchi et al., 2013). T1 weighted images were resliced (1 mm isotropic) and surface extraction and cortical parcellation for each individual structural image was carried out using Freesurfer version 5.1 (Fischl et al., 1999). The preprocessing followed the standard description available at (http://surfer.nmr.mgh.harvard.edu/). Image processing for morphometry was carried out blind to diagnosis. Cortical thickness was computed in accordance with the standard processing routines using Freesurfer with no control-point editing being required for this dataset (McCarthy et al., 2015). For this study, we employed Yeo’s Functional Atlas (17-networks) to parcellate the entire cortical surface (Yeo et al., 2011). Medial wall of the reconstructed hemispheres near the ventricles, proximal to the corpus callosum, was not included. The mean thickness of all vertices that were confined within the boundaries of the parcellated regions provided the network’s thickness value, averaged across the 2 hemispheres. To compute the index for spatial coherence, we calculated the standard deviation of vertex wise thickness within each network, expressed as a ratio of the mean thickness (coefficient of variation of thickness, CVT). Higher CVT of a network in a subject indicates lower spatial coherence within the network for that individual. This computation is described in Figure 1. Thus, we obtained 17 mean thickness and CVT values for further analysis. A template identifying these networks is shown in Supplemental Figure 1.

**Figure 1:**
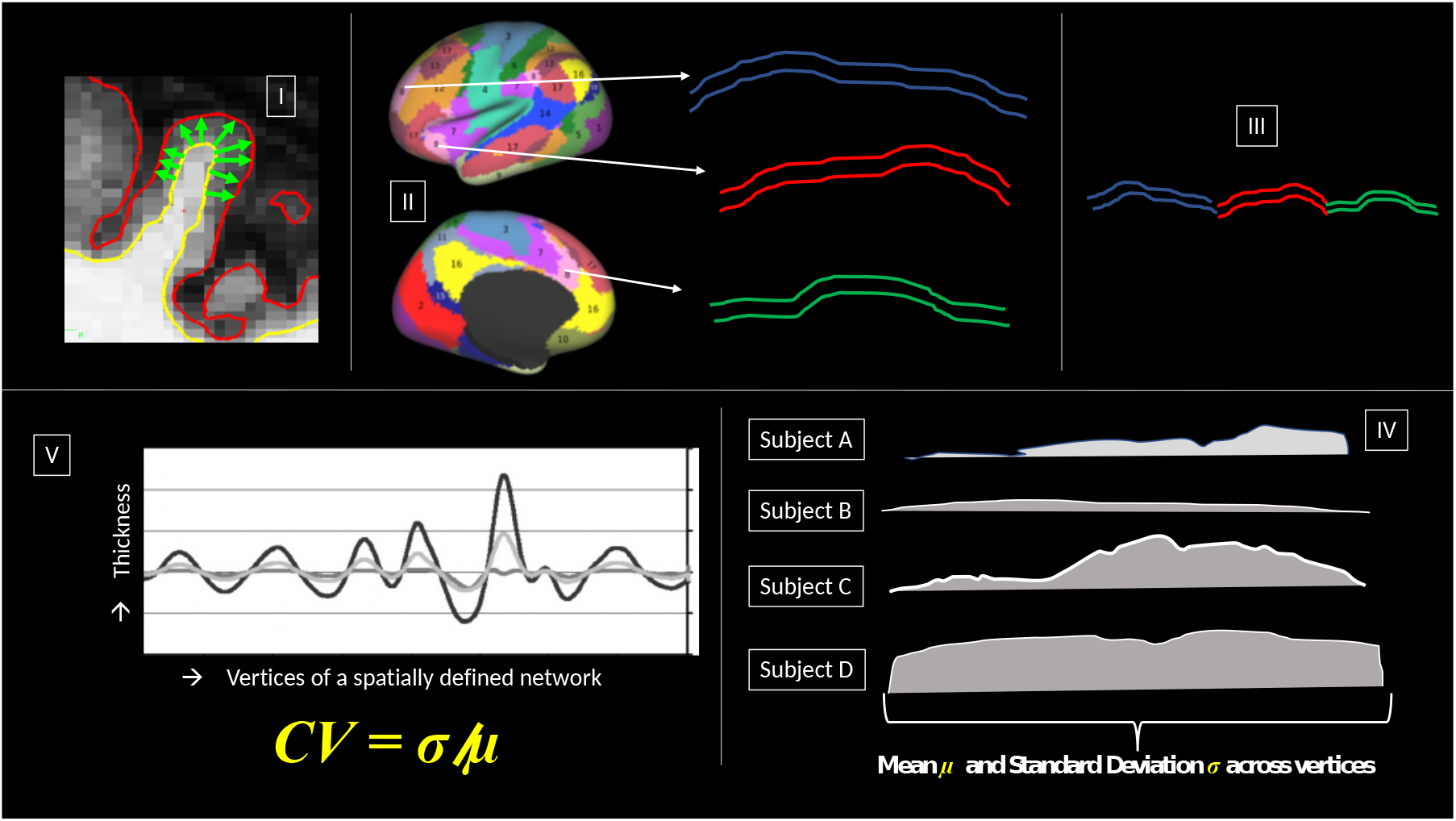
Estimating thickness-based spatial coherence within a functional network. I. For each vertex on grey-white boundary, the tangential distance to pial boundary provides a vertex-specific thickness measure, extracted using Freesurfer-based surface reconstruction. II. Using Yeo’s 17 functional networks, the anatomical boundaries are defined for cortical networks. For any given network (Salience Network in this example), included regions are spatially distributed, with thickness varying across the vertices. III. Vertices that make up each network show variable thickness, that can be averaged to obtain mean thickness of the functional network, while standard deviation across the vertices provides the measure of variability. Disruptions in the coordinated maturation and/or functional co-activation within these networks may result in increased variability in thickness across the network IV. Hypothetical vertexwise thickness values plotted in a spatial continuum for 4 subjects. Subject A shows moderate degree of variability with average mean thickness; Subject B shows low variability as well as low mean thickness; Subject C shows high degree of variability with high mean thickness; Subject D shows low variability but high mean thickness. V. As variability scales with mean values (Taylor’s law), the spatial coherence within a network is best captured by the coefficient of variation (CV = mean/standard deviation).

### Statistical Analysis

All statistical tests were carried out using SPSS version 25.0 (IBM Corp., Armonk, NY). T tests, Mann–Whitney U tests for non-normal data and chi-square tests for proportions were used to compare clinical and demographic variables between patients and controls. Independent 2-tailed t-tests were used to compare CVT and mean cortical thickness of 17 networks between patients and healthy controls with False Discovery Rate (FDR) corrected p<0.05 considered as statistically significant for the 34 comparisons that were made. We then used nonparametric correlation (Kendall’s rank correlation coefficient tau (Kendall, 1938)) to relate the TLI positive FTD scores to the CVT and mean thickness values of the networks with diagnostic differences, with FDR correction applied for the multiple correlations.

When undertaking a correlation with symptom severity, we assume that the scores vary continuously with brain structure. But structural changes may relate to the discrete presence and not vary continuously with severity. To ensure that our interpretations are not biased by this assumption, we divided the patient sample into binary categories based on the presence of positive FTD (pFTD+ vs pFTD-) defined as scores of >0.25 summed positive FTD score (disorganization), in line with the original work on TLI. The CVT and mean thickness values of the networks with diagnostic differences were directly compared between pFTD+ and pFTD-groups.

To test the specificity of our findings to positive FTD, we also repeated the above analyses with negative FTD scores. We also removed the variance in grey matter thickness and CVT explained by current antipsychotic defined daily dose (DDD), age, IQ, global functioning (GAF scores), and total intracranial volume and related the residuals to positive and negative FTD to establish specificity of the observed relationships.

## Results

Demographic and clinical features of the sample are presented in Table 1, highlighting match for age, gender and parental socioeconomic status between the 2 groups.

**Table 1:**
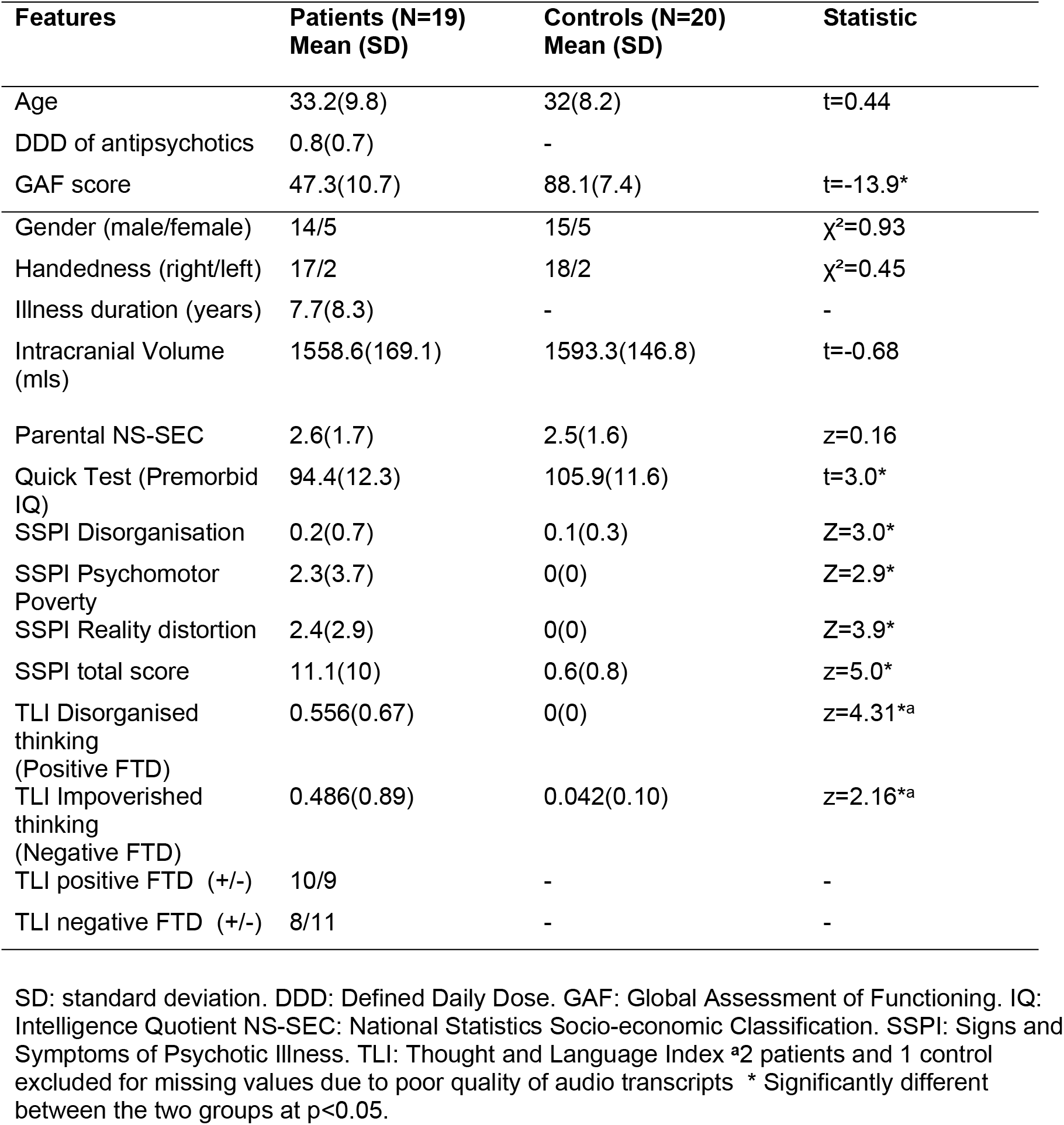
Clinical and demographic features

| Features | Patients (N=19)<br>Mean (SD) | Controls (N=20)<br>Mean (SD) | Statistic |
| --- | --- | --- | --- |
| Age | 33.2(9.8) | 32(8.2) | t=0.44 |
| DDD of antipsychotics | 0.8(0.7) | - |  |
| GAF score | 47.3(10.7) | 88.1(7.4) | t=-13.9* |
| Gender (male/female) | 14/5 | 15/5 | $\chi^2=0.93$ |
| Handedness (right/left) | 17/2 | 18/2 | $\chi^2=0.45$ |
| Illness duration (years) | 7.7(8.3) | - | - |
| Intracranial Volume (mls) | 1558.6(169.1) | 1593.3(146.8) | t=-0.68 |
| Parental NS-SEC | 2.6(1.7) | 2.5(1.6) | z=0.16 |
| Quick Test (Premorbid IQ) | 94.4(12.3) | 105.9(11.6) | t=3.0* |
| SSPI Disorganisation | 0.2(0.7) | 0.1(0.3) | Z=3.0* |
| SSPI Psychomotor Poverty | 2.3(3.7) | 0(0) | Z=2.9* |
| SSPI Reality distortion | 2.4(2.9) | 0(0) | Z=3.9* |
| SSPI total score | 11.1(10) | 0.6(0.8) | z=5.0* |
| TLI Disorganised thinking (Positive FTD) | 0.556(0.67) | 0(0) | z=4.31<sup>a</sup> |
| TLI Impoverished thinking (Negative FTD) | 0.486(0.89) | 0.042(0.10) | z=2.16<sup>a</sup> |
| TLI positive FTD (+/-) | 10/9 | - | - |
| TLI negative FTD (+/-) | 8/11 | - | - |

SD: standard deviation. DDD: Defined Daily Dose. GAF: Global Assessment of Functioning. IQ: Intelligence Quotient NS-SEC: National Statistics Socio-economic Classification. SSPI: Signs and Symptoms of Psychotic Illness. TLI: Thought and Language Index <sup>a</sup>2 patients and 1 control excluded for missing values due to poor quality of audio transcripts \* Significantly different between the two groups at p<0.05.

### Diagnostic differences

Network-based thickness differences between patients and healthy controls are shown in Table 2 (with descriptive labels as per Baker et al., 2014). The frontoparietal network and the midline default mode network showed the most significant reduction in cortical thickness (networks #12 and #16 of Yeo atlas) in patients when compared to controls. The cingulo-insular salience network and the frontoparietal networks (#8 and #12) had higher CVT in patients compared to controls, indicating spatial incoherence within these cognitive control networks in patients with schizophrenia.

**Table 2:**
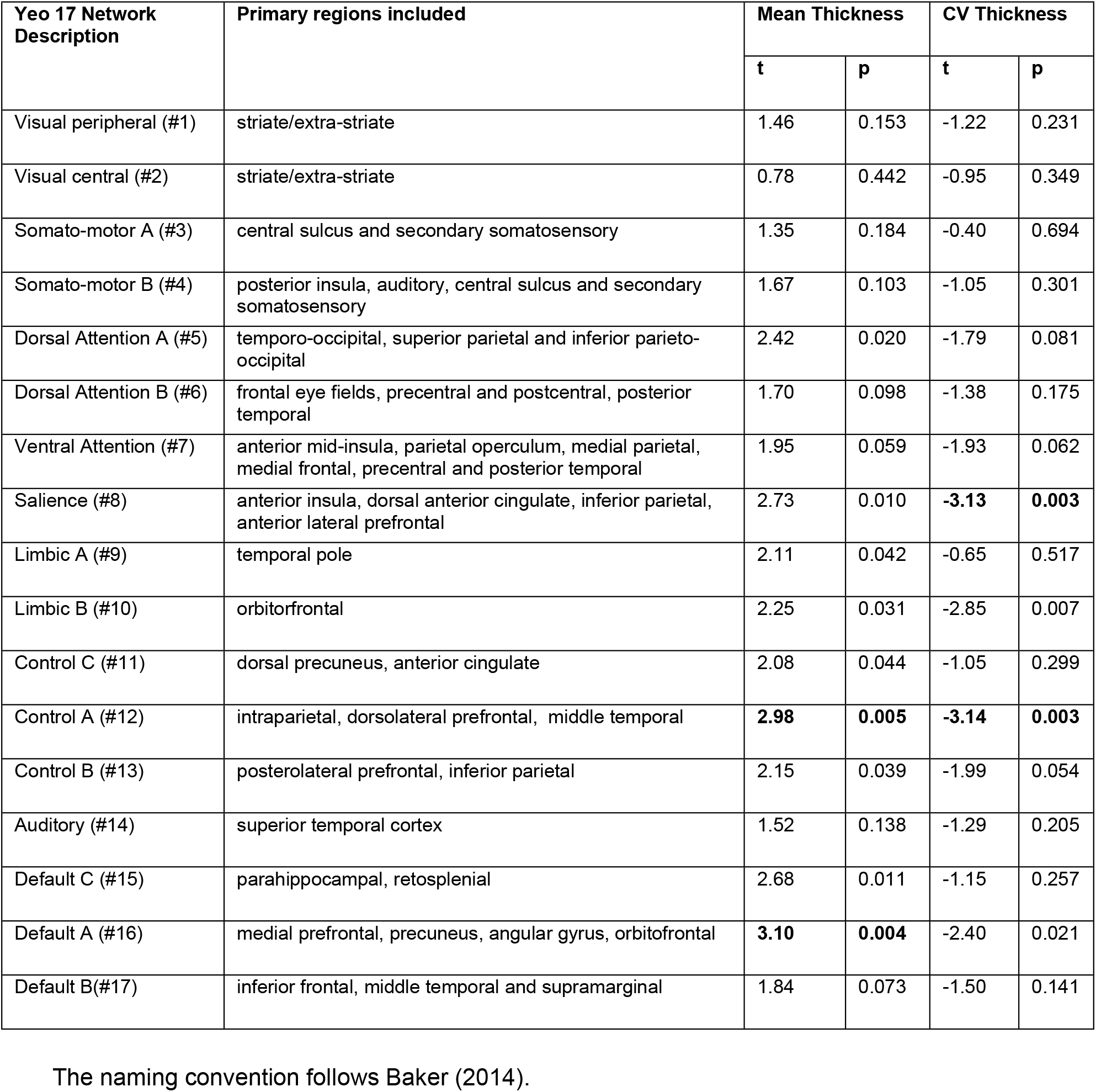
Group differences in mean thickness and spatial coherence

| Yeo 17 Network Description | Primary regions included | Mean Thickness |  | CV Thickness |  |
| --- | --- | --- | --- | --- | --- |
|  |  | t | p | t | p |
| Visual peripheral (#1) | striate/extra-striate | 1.46 | 0.153 | -1.22 | 0.231 |
| Visual central (#2) | striate/extra-striate | 0.78 | 0.442 | -0.95 | 0.349 |
| Somato-motor A (#3) | central sulcus and secondary somatosensory | 1.35 | 0.184 | -0.40 | 0.694 |
| Somato-motor B (#4) | posterior insula, auditory, central sulcus and secondary somatosensory | 1.67 | 0.103 | -1.05 | 0.301 |
| Dorsal Attention A (#5) | temporo-occipital, superior parietal and inferior parieto-occipital | 2.42 | 0.020 | -1.79 | 0.081 |
| Dorsal Attention B (#6) | frontal eye fields, precentral and postcentral, posterior temporal | 1.70 | 0.098 | -1.38 | 0.175 |
| Ventral Attention (#7) | anterior mid-insula, parietal operculum, medial parietal, medial frontal, precentral and posterior temporal | 1.95 | 0.059 | -1.93 | 0.062 |
| Salience (#8) | anterior insula, dorsal anterior cingulate, inferior parietal, anterior lateral prefrontal | 2.73 | 0.010 | **-3.13** | **0.003** |
| Limbic A (#9) | temporal pole | 2.11 | 0.042 | -0.65 | 0.517 |
| Limbic B (#10) | orbitofrontal | 2.25 | 0.031 | -2.85 | 0.007 |
| Control C (#11) | dorsal precuneus, anterior cingulate | 2.08 | 0.044 | -1.05 | 0.299 |
| Control A (#12) | intraparietal, dorsolateral prefrontal, middle temporal | **2.98** | **0.005** | **-3.14** | **0.003** |
| Control B (#13) | posterolateral prefrontal, inferior parietal | 2.15 | 0.039 | -1.99 | 0.054 |
| Auditory (#14) | superior temporal cortex | 1.52 | 0.138 | -1.29 | 0.205 |
| Default C (#15) | parahippocampal, retrosplenial | 2.68 | 0.011 | -1.15 | 0.257 |
| Default A (#16) | medial prefrontal, precuneus, angular gyrus, orbitofrontal | **3.10** | **0.004** | -2.40 | 0.021 |
| Default B (#17) | inferior frontal, middle temporal and supramarginal | 1.84 | 0.073 | -1.50 | 0.141 |

The naming convention follows Baker (2014).

### Relationship with positive FTD

A higher CVT of the salience network (tau = 0.41 p = 0.024) and lower mean thickness in frontoparietal network (tau = -0.498, p = 0.006) predicted the severity of positive FTD (after FDR correction). CVT of the frontoparietal network (tau = 0.298, p = 0.10) and mean thickness of the default mode network (tau = -0.355, p = 0.05) did not show a significant relationship with the severity of positive FTD, even at the uncorrected level.

Residuals adjusted for the linear influence of antipsychotic dose, age, IQ, overall functioning, and total intracranial volume continued to demonstrate a significant relationship between higher CVT of the salience network (tau = 0.37 p = 0.04; Figure 2) and lower mean thickness in frontoparietal control network (tau = -0.48, p = 0.008) with the severity of positive FTD. Positive FTD did not relate to the adjusted residuals of other structural measures involving the default mode and the language networks.

**Figure 2:**
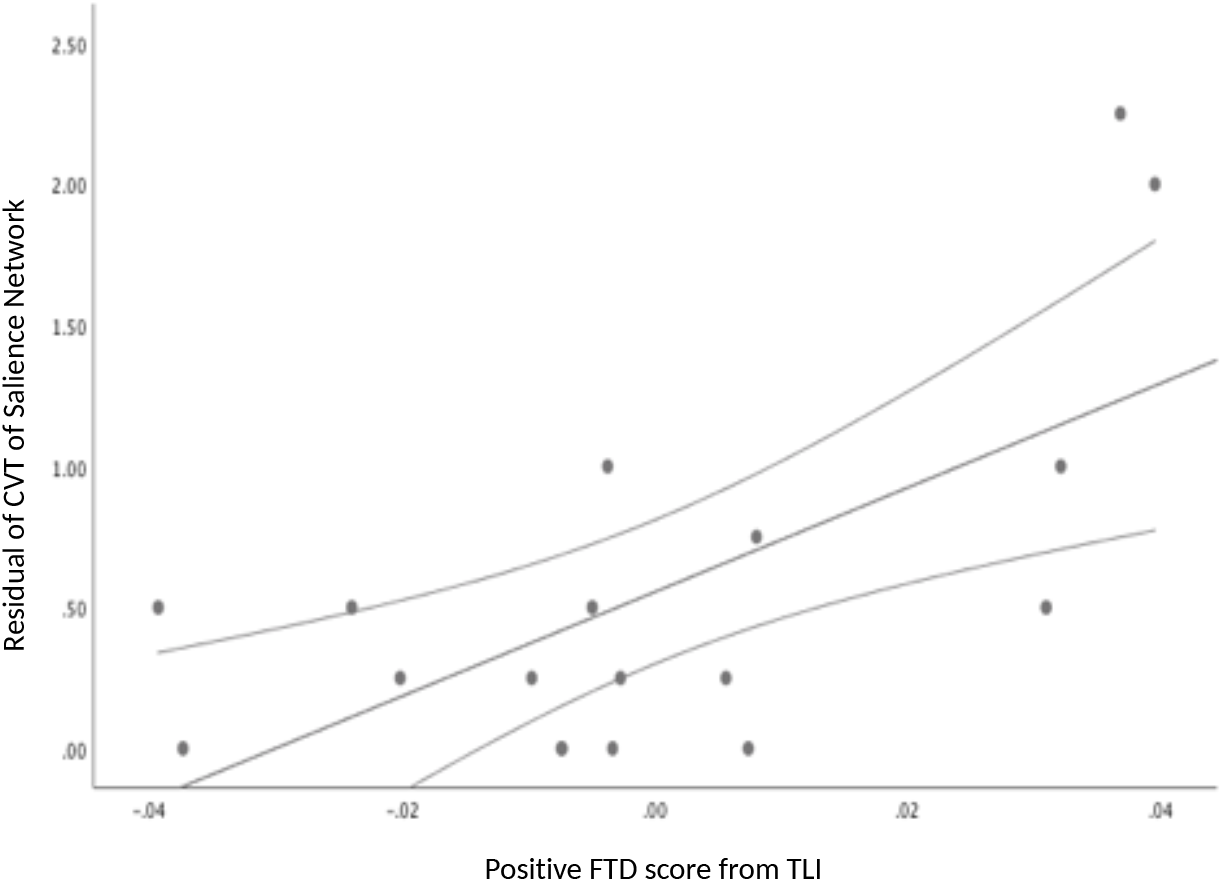
Relationship between spatial incoherence of the Salience Network and positive formal thought disorder. The residuals linearly adjusted for age, gender, intracranial volume, IQ, global functioning and antipsychotic dose are plotted on y-axis, and summed scores of positive formal thought disorder on the x-axis. Note that the scatter-plot depicts the linear relationship, while non-parametric correlations were used in the statistical analysis. CVT: Coefficient of Variation of Thickness; FTD: Formal Thought disorder; TLI: Thought & Language Index.

In direct group comparison, pFTD+ group showed significant reduction in mean thickness of the frontoparietal control network (t = 2.93, p=0.009) and increase in spatial incoherence indexed by CVT of the salience network (t=-2.96, p=0.009), indicating that both the presence and the severity of positive FTD is influenced by these 2 networks.

### FTD and the structure of language networks

We did not find significant correlations between the mean thickness and CVT of the 2 language networks (#14 and 17 of Yeo atlas) that include Wernicke’s and Broca’s regions and positive FTD scores, even at an uncorrected level (absolute values of tau =0.18 to 0.30; p= 0.06-0.31). Nevertheless, pFTD+ group showed a significant reduction in mean thickness of the auditory network that included superior temporal cortex (#14) (t = 3.68, p=0.002) when compared to the pFTD-group, indicating that the presence of positive FTD, rather than its severity, is influenced by the structure of the language network comprising of superior temporal gyrus.

Furthermore, we did not find significant correlations between the thickness and CVT of the frontoparietal network, CVT of the salience network, the thickness of default network or the mean thickness and CVT of the 2 language networks (#14 and 17) with negative FTD scores, even at an uncorrected level (absolute values of tau = 0.07-0.32; p= 0.09-0.71). On binary analysis, nFTD+ group showed a significant reduction in mean thickness of the network that included middle temporal gyrus (#17) (t = 2.3, p=0.034) when compared to the nFTD-group, but this did not survive FDR correction for the t tests involving the language networks. Adjusting for the linear influence of antipsychotic dose, age, IQ, overall functioning, and total intracranial volume did not alter these results.

## Discussion

To our knowledge, this is the first study investigating cortical thickness of functional networks in relation to FTD in schizophrenia. In this study, we have shown that (1) clinically stable and medicated individuals with schizophrenia exhibit reduced thickness of the frontoparietal and default mode networks, and spatial incoherence affecting the salience and the frontoparietal network (2) Patients with more severe burden of positive FTD had reduced frontoparietal network thickness and spatial coherence of the salience network (3) The presence of positive FTD, but not its severity, related to the reduced thickness of the auditory network comprising of the superior temporal cortex. Taken together, these results indicate that the severity of FTD is contingent upon the degree to which domain-general control aspects of information processing (i.e. frontoparietal and salience systems (Cole et al., 2014; Fedorenko et al., 2013)) are affected in schizophrenia, over and above the structural deficits in the language areas (i.e. temporal cortex).

While the reduced thickness of the frontoparietal and default mode regions has been reported consistently in many previous studies of schizophrenia (Ehrlich et al., 2012; Heinrichs et al., 2017; Park et al., 2018; Shafiei et al., 2019), the presence of spatial incoherence has not been investigated so far to our knowledge. The presence of higher than expected variation in the vertexwise thickness of large-scale cortical networks involved in information processing (salience and frontoparietal cognitive control) may indicate the possibility of a disturbance in coordinated structural maturation (Alexander-Bloch et al., 2013b). A developmental lag in intracortical myelination or synaptic pruning (Fjell et al., 2015) of one region relative to the others within a network can result in such incoherence. Given the genetic control of intracortical myelination of during adolescence is closely related to the risk of schizophrenia (Whitaker et al., 2016), this possibility requires further investigation. It is also possible that the harmonious functional deployment of the network nodes do not occur as expected in patients, leading to a structural reconfiguration indicated by increased vertex-wise variance in thickness. This possibility has been raised previously in a study of spatial topography of functional connectivity in schizophrenia (Skudlarski et al., n.d.). While the results presented here cannot distinguish these two mechanisms, our results indicate that within-subject structural variability is an useful source of information to understand the elusive neural basis of complex symptoms such as formal thought disorder.

Our results partially confirm the long suspected role of language network in positive FTD, and add further clarity to the putative mechanistic basis of the observed disorganized thinking in schizophrenia. While structural changes involving both the cognitive control (frontoparietal and salience) and the language (auditory) network are seen in patients with positive FTD, the severity of the clinically expressed FTD relies on the degree to which the control systems are affected. In particular, both a reduction in the amount of grey matter tissue in the frontoparietal network and the variable distribution of grey matter within the salience network, appear to mediate the expressed severity of the disorganized thought in schizophrenia. This is consistent with the previously reported relationship between disorganisation and the disturbed functional relationships between the nodes of the salience network (ACC and insula)(Liddle et al., 1992), as well as a recent observation that the functional flexibility of the salience network predicts conceptual disorganisation but not the negative symptoms (Supekar et al., 2019). The notion of ‘cognitive control systems’ tuning the expression of a language system abnormality is in line with the current notions on cognitive control of language (Blank et al., 2014; Fedorenko and Varley, 2016), and consistent with the clinical observations that the severity of FTD increases when excessive cognitive demands (e.g. distraction, longer conversations) are placed on an individual with schizophrenia (Becker et al., 2012; Docherty, 2012).

In our previous study, we noted that a reduction in grey matter volume involving the insula, precuneus and lateral temporal regions in the presence of an increase in grey matter volume involving the cingulate and lateral prefrontal regions predicted higher burden of negative FTD, but not the positive FTD (Palaniyappan et al., 2015). Our current observations relate positive FTD to the salience (insula, ACC) and lateral frontoparietal as well as the auditory network (lateral temporal) structure; such a relationship was not seen for negative FTD. The lack of relationship between cortical thickness and negative FTD is in line with one previous report (Winkelbeiner et al., 2018), and indicate that maturational defects that affect morphometric properties other than thickness (e.g. surface area or gyrification (Palaniyappan and Liddle, 2012)) may be more critical for developing negative FTD. Taken together, these results indicate that both forms of persistent FTD result from cross-network structural aberrations involving both the language-specific as well as cognitive control systems.

There are several limitations in our study. First, we had a limited sample size that precluded us from undertaking a mass univariate approach to the analysis. Nevertheless, it is important to note that we had sufficient power to reject the 2 primary null hypotheses (no patient vs. control differences and lack of structural correlates of positive FTD) with a low probability of false discovery. The possibility of type 2 error due to small sample cannot be fully dismissed for the null results relating to the language network and secondary analysis of the negative FTD scores. We urge caution when interpreting these negative results. Second, unlike Sans-Sansa et al. (2013), our patient sample was selected for its clinical stability but not for exhibiting FTD. While this may be a limitation, this enabled us to capture the milder but persistent aspects of FTD and thus provided a more representative sample when seeking morphological correlates of persistent as opposed to acute FTD. Third, we did not measure the subjective component of thought disorder that can be quantified using more recent instruments such as the Thought and Language Disorder (TALD) scale (Kircher et al., 2014). Nevertheless, most of the existing FTD-specific instruments consistently differentiate the positive and negative FTD dimensions, indicating that the biological mechanisms discussed here are likely to be valid irrespective of the specific instrument employed.

In summary, we demonstrate that the cortical thickness of both cognitive control and language networks underlie the positive FTD in schizophrenia. The structural integrity of cognitive control networks is a critical determinant of the expressed severity of persistent FTD in schizophrenia.

## Data Availability

Reasonable requests for the anonymised data reported in this manuscript can be made to the corresponding author

---

**Figure S1:**
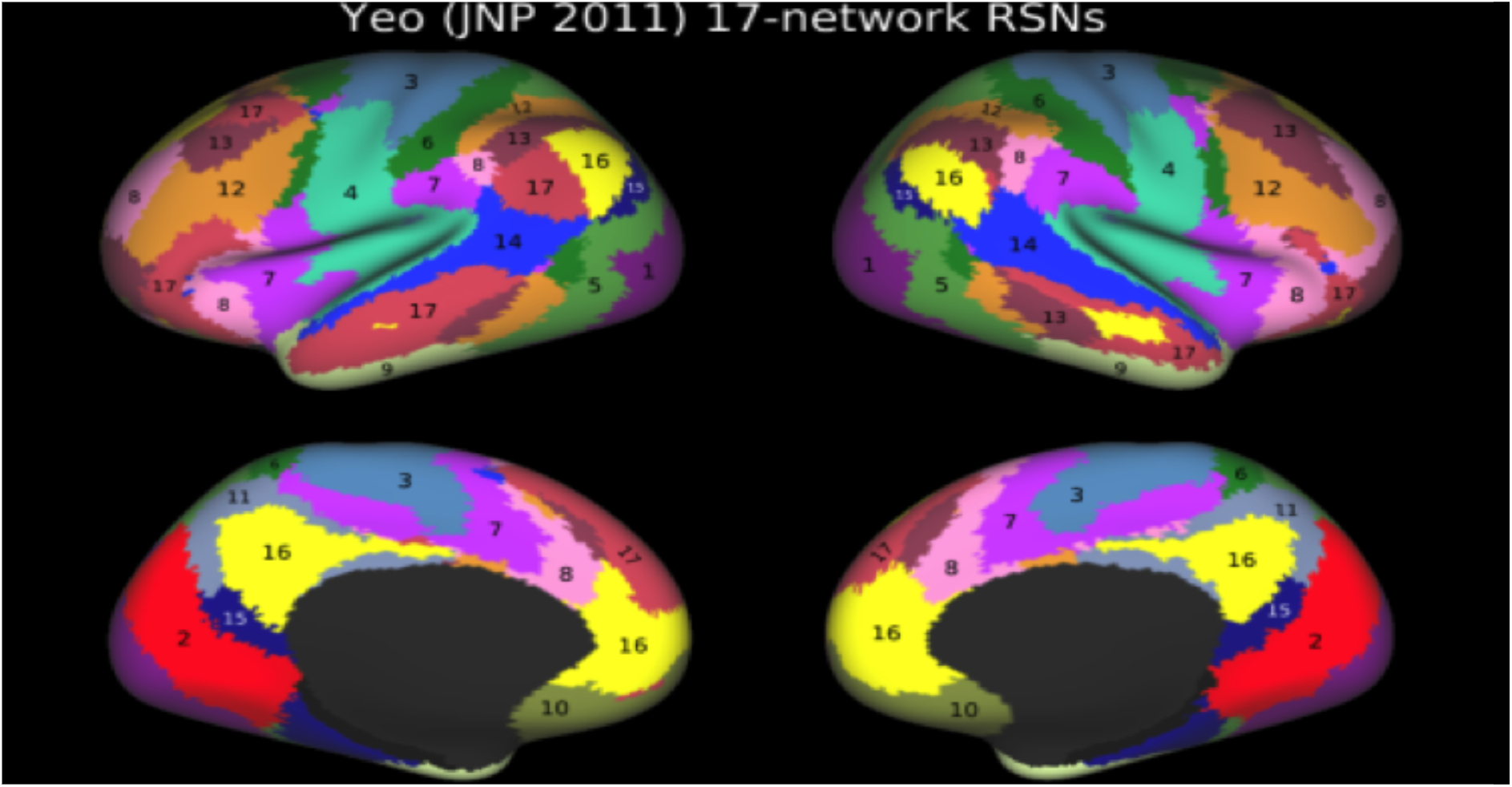
Yeo 17 network parcellations. This figure is obtained from https://balsa.wustl.edu/W8wK. The labels are based on the original description of Yeo’s 17-netorks parcellation provided at https://surfer.nmr.mgh.harvard.edu/fswiki/CorticalParcellation_Yeo2011. The naming convention and regional description as per Baker (2014) is displayed in Table 2.

## Cortical thickness and formal thought disorder in schizophrenia

### Ethics

All subjects were recruited from the county of Nottinghamshire, UK with informed consent to participate obtained in accordance with the approval from the National Research Ethics Committee, Nottinghamshire. NHS REC Ref: 10/H0406/49

